# Efficacy and safety of a drug-free, barrier-forming nasal spray for allergic rhinitis: randomized, open-label, crossover noninferiority trial

**DOI:** 10.1101/2021.08.02.21261374

**Authors:** Julian Nehrig, Nicole Grosse, Ilja P. Hohenfeld, Fabio Fais, Thomas Meyer, Jens M. Hohlfeld, Philipp Badorrek

**Affiliations:** Fraunhofer ITEM, Feodor-Lynen-Str. 15, 30625 Hannover, Germany; Altamira Medica AG, Bahnhofstrasse 21, 6300 Zug, Switzerland; Department of Respiratory Medicine, Hannover Medical School, Hannover, Germany; German Center for Lung Research (DZL), BREATH, Hannover, Germany

**Keywords:** Allergen challenge, Allergy, Bentonite, *Dactylis glomerata*, Hydroxypropyl methylcellulose, Medical device, Nasal spray, Pollen, Rhinorrhea, Thixotropy

## Abstract

**Background:** Symptoms of allergic rhinitis can be reduced by nonpharmacological nasal sprays that create a barrier between allergens and the nasal mucosa. A new nasal spray (AM-301) containing the clay mineral bentonite was tested for its ability to reduce symptoms to grass pollen.

**Methods:** This open label, crossover, noninferiority trial compared the efficacy and safety of AM-301 to that of hydroxypropyl methylcellulose (HPMC; Nasaleze Allergy Blocker), an established barrier method. Adults with seasonal allergic rhinitis were exposed to *Dactylis glomerata* pollen, in the Fraunhofer Allergen Challenge Chamber, first without protection and then protected by HPMC or AM-301 (7 days apart). Efficacy was assessed from total nasal symptom score (TNSS), nasal secretion weight, and subjective rating. The primary endpoint was the difference, between AM-301 and HPMC, in least square mean change in TNSS over a 4-hour exposure to allergen.

**Results:** The study enrolled 36 persons, and 35 completed all study visits. The mean TNSS was 5.91 during unprotected exposure, 5.20 during protection with HPMC, and 4.82 during protection with AM-301. The difference in least square means between the two treatments was -0.39 (95% CI, -0.89 to 0.10), establishing the noninferiority of AM-301. No difference in mean weight of nasal secretions was observed between the treatments. Efficacy was rated as good or very good for AM-301 by 31% and for HPMC by 14% of subjects. 16 subjects reported adverse events with a relationship to AM-301 or HPMC; most adverse events were mild and none was serious.

**Conclusions:** AM-301 demonstrated noninferiority towards HPMC in the primary endpoint and was perceived better in subjective secondary endpoints. Both barrier-forming products had a persisting protective effect over 4 hours and were safe.

**Trial registration:** German Clinical Trial Register (DRKS00024356). EUDAMED (CIV-20-10-034870).

## Background

Seasonal allergic rhinitis (SAR) has a significant impact on quality of life [1–3]. Its high prevalence in Europe (e.g. 24.4% [95% CI, 16.5% to 24.6%] in Germany and 24.5% [95% CI, 21.0% to 28.0%] in France) [4] establishes the need to continue to develop effective and safe therapies. Moreover, because patients with SAR are at risk of asthma [5] they would benefit from reliable protection from allergen exposure.

Several classes of treatment for SAR are currently available. Oral H1 antihistamines are effective treatments [6]. A newer generation of these drugs, including desloratadine and cetirizine, have less side effects (e.g. sedation and drowsiness) and eliminated the safety concerns of the first generation [7], resulting in a strong recommendation for their use in SAR treatment [1]. Nevertheless, side effects remain problematic for a small proportion of patients (reviewed in [8]), and the incidence of somnolence can be as high as 13% for subjects taking cetirizine[9]. Another class of drugs used as first-line SAR therapy is intranasal corticosteroids. These drugs are more effective than oral antihistamines in reducing nasal and ocular symptoms [1]. However, side effects such as epistaxis [10] limit the acceptability of intranasal corticosteroids. SAR can also be treated effectively with allergen immunotherapy, but this therapy requires high patient compliance for regular clinical visits [11] and its benefits may diminish over time [1]. Therefore, there remains a need for new approaches to SAR symptom relief.

One promising approach to SAR prevention consists of blocking any contact between allergen and nasal mucosa [12]. These “barrier” approaches may be used as complements to drug therapies or as standalone methods for patients who do not accept or tolerate other treatments. The effectiveness of a barrier between allergen and airway was exemplified by the reduction of allergic rhinitis symptoms during the COVID-19 pandemic by the frequent use of face masks [13]. In addition to masks, which are applied externally, several intranasal barriers are available. Among these, topical application of hydroxypropyl methylcellulose (HPMC) powder has been established as both effective and safe [14].

A new barrier method against airborne allergens is the nasal spray AM-301 (marketed as Bentrio), a substance-based medical device. AM-301 is a thixotropic gel emulsion that is sprayed into the nostrils via a standard nasal spray applicator. Before use, the applicator bottle must be shaken briefly to reduce the viscosity of the formulation, which transitions from a gel to a liquid state and thus becomes sprayable. Then, when the liquid settles on the nasal mucosa, it creates a protective film as the formulation returns back to a gel state. These rheological properties are mediated by the interaction of the formulation’s key ingredient, the mineral bentonite, with the other ingredients. AM-301 contains no active pharmaceutical ingredient and exerts its protective properties through physical effects only.

This paper reports the results of a clinical trial on the safety and efficacy of the investigational medical device AM-301. In this noninferiority trial, AM-301 was compared with HPMC powder, considered an established barrier method. The study was conducted in the Fraunhofer Allergen Challenge Chamber (ACC), which creates a controlled environment for pollen exposure and is a validated method [15] that has been used successfully in several clinical studies [16–19].

## Material and methods

### Ethics statement and study design

This single-center, open-label, randomized crossover trial was conducted at the Fraunhofer Institute for Toxicology and Experimental Medicine (Fraunhofer ITEM, Hannover, Germany). The trial was performed between February and April 2021, before the expected start of the grass pollen season in Northern Germany [20,21] and Berlin [22]. The study protocol was approved by the Ethics Committee of Hannover Medical School prior to study start. The trial was registered in the German Clinical Trial Register (DRKS00024356) and with EUDAMED (CIV-20-10-034870). Because this study involved a class I medical device, it was granted a waiver for formal approval by the Federal Institute of Drugs and Medical Devices. All study participants provided written informed consent, including the consent for publication of the trial data in a scientific journal.

The study was conducted in the Fraunhofer ACC, which is an established, validated setting to create indoor grass pollen atmospheres for clinical trials [15]. Throughout each ACC visit, humidity, temperature, airflow and allergen load were monitored. For this trial, *Dactylis glomerata* pollen (Pharmallerga, Lisov, Czech Republic) was used as the allergen.

People with a diagnosis of SAR were initially eligible for this study if they were positive on a prick test to grass pollen and met the medical inclusion and exclusion criteria described below. Full eligibility was established if they were responsive to *Dactylis glomerata* pollen during an unprotected screening ACC visit, i.e. a 4-hour exposure to pollen in the ACC without any barrier method. Responsiveness was defined as achieving a total nasal symptom score (TNSS) ≥6 at least twice during the exposure. To eliminate any possible seasonal influence on the study outcomes, only subjects with a TNSS ≤3 before allergen exposure were allowed to enter the ACC; all those with TNSS >3 were excluded from testing. Fully eligible subjects were invited for two treatment ACC visits, i.e. additional 4-hour exposures in the ACC with application of a barrier-forming medical device (for an overview of study visits see **Figure 1**). The first treatment ACC visit was scheduled at least 7 days after the unprotected screening ACC visit, and the second treatment ACC visit was scheduled 7 days after the first. The subjects were randomized, based on a predefined randomization list, to two treatment sequences.

**Figure 1.**
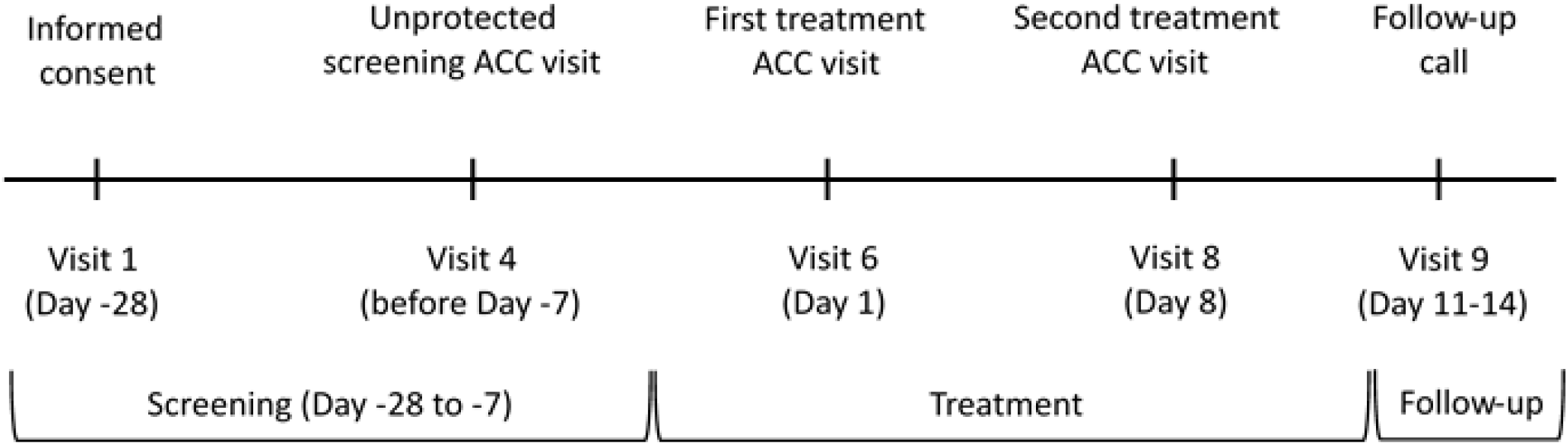
Study plan. The main visits for screening and treatment are indicated. Each ACC visit involved a 4-hour grass pollen exposure. At visits 2, 3, 5 and 7, tests for SARS-CoV-2 were performed. ACC, allergen challenge chamber.

Sequence A received AM-301 at the first treatment ACC visit and the comparator device Nasaleze at the second treatment ACC visit, while Sequence B received Nasaleze first and AM-301 successively. A study physician applied the medical device using a standardized procedure. Then, subjects entered the ACC within 10±2 minutes.

At each ACC visit, TNSS and nasal secretion weight were assessed every 20 minutes and every 60 minutes, respectively. The safety of subjects was monitored via vital signs and peak expiratory flow. Throughout the study, subjects underwent oropharyngeal swabbing to test for SARS-CoV-2 infection by PCR on separate visits. In addition, SARS-CoV-2 rapid antigen testing of nasopharyngeal swabs was performed 1 hour before entering the ACC. Any positive test result for SARS-CoV-2 led to exclusion from the study. A follow-up visit was performed several days after the second treatment ACC visit.

### Study population

The study enrolled men and women aged 18 to 65 years with a diagnosis of SAR to grass pollen documented by a positive skin prick test. Women of childbearing potential had to use a safe method of contraception throughout the duration of the study. Exclusion criteria were: a body mass index <18 or >32 kg/m^2^, asthma requiring medication other than inhaled short-acting ß2-agonists, an acute infection within 4 weeks prior to the start of the study, allergen-specific immunotherapy within 2 years prior to the study, an upper respiratory tract infection including rhinitis within 14 days before the trial, severe septum deviation or other structural nasal abnormalities that would prevent the use of a nasal spray, a history of drug or alcohol abuse in the past 2 years, and compromised lung function (FEV1 <80% or FEV1/FVC <70%). Other medical conditions with mild or controlled symptoms that would not interfere with the study outcome and would not put the subject at risk were tolerated. Use of paracetamol and short-acting ß2-agonist aerosols were allowed, while other allergy medications (e.g. inhaled or intranasal glucocorticoids and oral antihistamines) were not allowed within defined pre-trial wash-out periods or during the trial, with the exception of short-acting eye or nose drops (e.g. azelastine) used immediately after the allergen challenge. Other concomitant medication use was allowed as judged by the investigators.

### Primary and secondary endpoints

The primary endpoint of the study was the difference between groups treated with AM-301 or HPMC in least square mean change of TNSS over the 4-hour (240 min) exposure to allergen. Secondary endpoints included differences between AM-301 and HPMC treatments in mean changes in individual TNSS items, at individual time points of TNSS assessment, in mean changes relative to the unprotected screening ACC visit, in nasal secretion weights, and in the overall ratings of efficacy and tolerability by both the subjects and the investigators. Furthermore, adverse events and peak expiratory flow after each challenge were evaluated. Secondary endpoints were analyzed in an exploratory approach.

### Efficacy assessments

Total nasal symptom score (TNSS) was determined using a four-item questionnaire that investigates nasal congestion, rhinorrhea, nasal itching, and sneezing [23]. Each symptom is scored on a 4-point severity scale (0, none; 1, mild; 2, moderate; 3, severe), and TNSS is the sum of each score, for a maximum value of 12. Subjects received an explanation of items and of the overall scale (in German) before the first assessment. TNSS was assessed before entering the ACC (baseline TNSS) and every 20 min after challenge start (13 time points in total).

Nasal secretion weight was measured by an established method [24,25]. Briefly, each subject was given four pre-weighed packs of tissues in collection bags, i.e. one bag with packs of tissues for every hour in the ACC. After every hour, the bag with used and unused tissues was collected and weighed. The increase in weight [g] during an hour of allergen exposure was considered due to rhinorrhea.

Subjective ratings of efficacy and tolerability were provided by both the subjects and the investigators after ACC visits with device application. Efficacy and tolerability were rated on an ad hoc questionnaire with a 4-point scale (very good, good, moderate, poor).

### Medical devices tested in the study

The marketed form of HPMC powder used as the comparator in this study was Nasaleze Allergy Blocker (Nasaleze International, Douglas, Isle of Man). It does not contain pharmaceutically active compounds and is administered by insufflation into the nose using a proprietary spray bottle. It also contains mint powder. One puff of Nasaleze was administered per nostril.

The investigational medical device, AM-301 (marketed as Bentrio; Altamira Medica, Zug, Switzerland), is a nasal spray designed to form a protective barrier on the nasal mucosa. The ingredients for this aqueous-based gel emulsion are all generally recognized as safe. The key component, bentonite, is an absorbent aluminum phyllosilicate clay consisting mostly of montmorillonite. Additional information about the composition and in vitro safety and efficacy of AM-301 is available in a companion publication [26]. AM-301 was provided in glass bottles with metered-dose spray pumps. One application of 0.14 mL was administered per nostril.

### Safety procedures

Before and after ACC visits, subjects were monitored for vital signs by noninvasive blood pressure and heart rate recordings. At each visit, subjects reported any and all adverse events, which were then assessed by a study physician as mild, moderate or severe. During the challenge, every 2 hours until bedtime, and once the next morning, subjects monitored their lung function using a peak flow meter (Vitalograph, Hamburg, Germany) to measure peak expiratory flow (PEF). Salbutamol was provided to every subject after ACC as rescue medication, and subjects were instructed to use it if PEF dropped by more than 20% compared to before allergen challenge.

### Statistical analysis

Statistical analysis was performed using Proc Mixed in SAS (SAS Institute, Cary, USA). To analyze differences in mean change in TNSS averaged over 240 min (the primary endpoint), a linear mixed model with fixed effect terms was fitted for the sequence (Sequence A: first AM-301, then HPMC; Sequence B: first HPMC, then AM-301), the period (effect between ACC visits regardless of the sequence), and the treatment (application of AM-301 vs. HPMC). From the model, least square means (LS_Means_) and corresponding two-sided 95% confidence intervals (CI) were extracted. The pre-challenge value of TNSS (baseline TNSS) was subtracted from the values obtained during each challenge per subject. Baseline TNSS was included in the statistical analyses as a covariate.

The model was also used for predefined exploratory analyses of the secondary endpoints. Briefly, mean change in TNSS at individual time points (20-min intervals until 240 min) and in individual TNSS items (nasal congestion, rhinorrhea, nasal itching, and sneezing) were analyzed using the model with the respective dependent variables. The model was adapted to evaluate the overall device effect on TNSS (unprotected screening ACC visit vs. treatment ACC visit with AM-301, and unprotected screening ACC visit vs. treatment ACC visit with HPMC) by replacing the covariate baseline TNSS values with mean TNSS values of the unprotected screening ACC visit as covariate. Nasal secretion weight was analyzed using this adapted model with the respective dependent variable.

### Sample size calculation for noninferiority

The study investigated the noninferiority of AM-301 towards HPMC and was powered according to this hypothesis. Therefore, the primary endpoint translates to ***μ***_***HP***_ ***− μ***_***AM***_ ***>*** − Δ, where ***μ***_***AM***_ and ***μ***_***HP***_ represent the true mean changes from baseline TNSS of AM-301 and HPMC, respectively, and Δ is the minimum clinically important difference (MCID). The MCID was defined as a 1.2-point difference in the TNSS. This is slightly above the 1-point difference suggested for use in ACC studies of human dust mite allergy [27] and in grass pollen-induced allergic rhinoconjunctivitis [28] (both studies used a 4-item nasal symptom score similar to the TNSS). Prior studies at Fraunhofer ITEM [16, 29] provided within-subject TNSS data over 120 min following grass pollen exposure in the Fraunhofer ACC: when data from these two studies were combined, the SD of the TNSS change from baseline was estimated to be 2.2 (90% CI, 2.1 to 2.5). With an MCID of 1.2 points and an SD of 2.2, an effect size of 0.55 was calculated, which is considered moderate (according to Cohen’s rule of thumb). With 32 subjects intended for randomization (16 per sequence), the study had 84% power at the 1-sided alpha level of 0.025 to reject the null hypothesis when it is false. To account for possible dropouts, the planned sample size was increased to 36 subjects.

## Results

For this noninferiority trial of AM-301 compared with HPMC powder, 72 subjects with SAR were screened for eligibility, and 36 subjects who matched all the inclusion and exclusion criteria were included and randomized (**Figure 2**). Overall, 35 subjects completed all visits of the clinical investigation, while one subject withdrew after the first treatment ACC visit and did not receive AM-301. Results reported here refer to the intention to treat (ITT) population with all 36 subjects.

**Figure 2.**
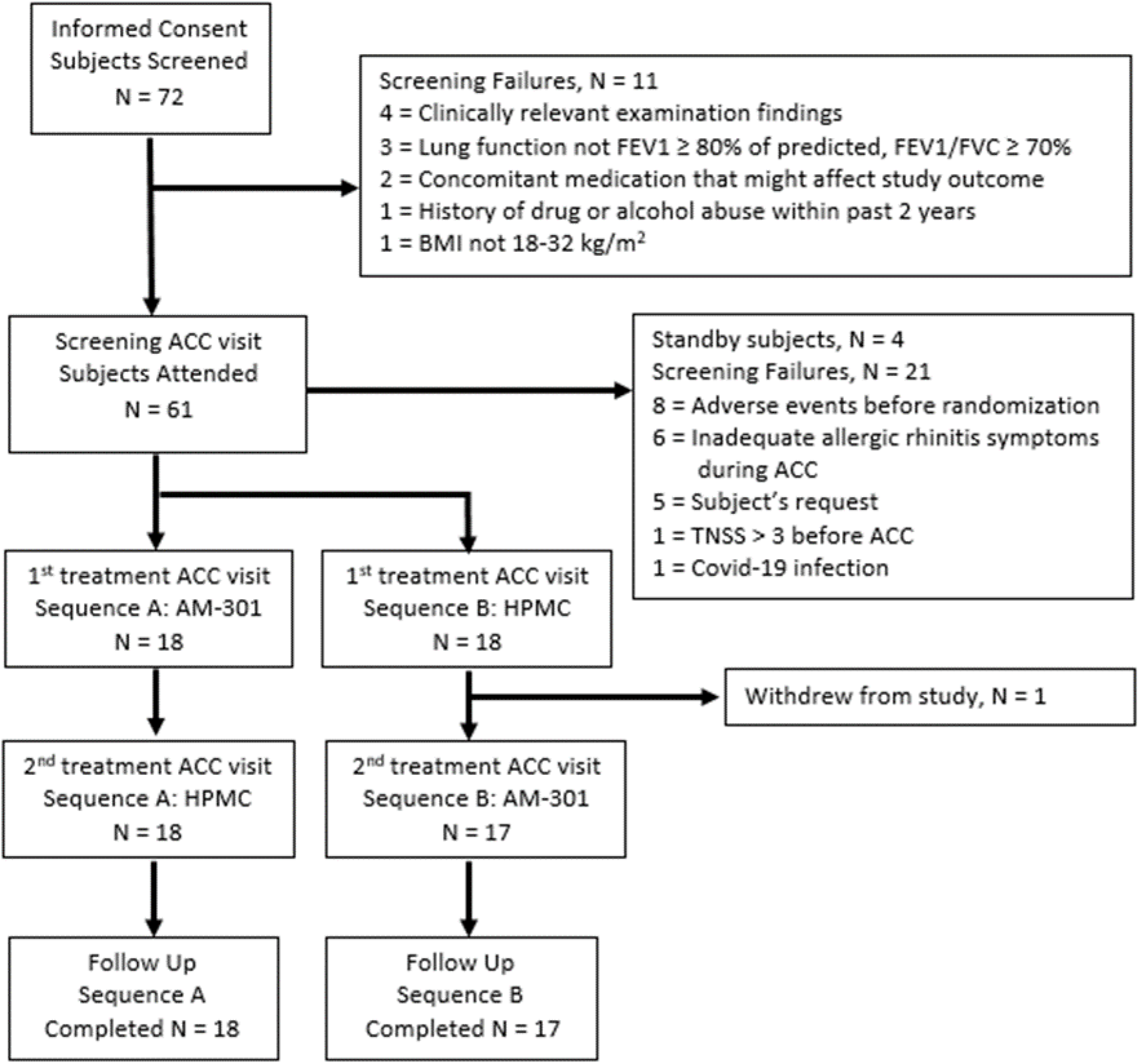
Subject disposition including reasons for exclusion. One subject withdrew from the study due to scheduling problems. ACC, allergen challenge chamber; AEs, adverse events; BMI, body mass index; FEV, forced expiratory volume; FVC, forced vital capacity; TNSS, total nasal symptom score

The study subjects had a mean age of 35.3 years (range, 21-62 years) (**Table 1**). All subjects were white and 25 (69%) were male. There were no study protocol deviations that could have impacted on patient safety or the outcome of TNSS and nasal secretion assessments.

**Table 1.** Clinical characteristics of the study subjects, according to the treatment sequence

| Characteristic |  | Sequence A<br>V6 AM-301; V8 HPMC<br>(N=18) | Sequence B<br>V6 HPMC; V8 AM-301<br>(N=18) | Overall<br>(N=36) |
| --- | --- | --- | --- | --- |
| Age (years), mean (SD) |  | 35.8 (12.4) | 34.8 (11.0) | 35.3 (11.6) |
| Gender, n (%) |  |  |  |  |
|  | Male | 13 (72) | 12 (67) | 25 (69) |
|  | Female | 5 (28) | 6 (33) | 11 (31) |
| Height (cm), mean (SD) |  | 176.9 (8.1) | 177.6 (8.2) | 177.2 (8.0) |
| Weight (kg), mean (SD) |  | 78.5 (12.6) | 81.6 (10.7) | 80.0 (11.6) |
| BMI (kg/m<sup>2</sup>), mean (SD) |  | 25.0 (3.2) | 25.8 (2.5) | 25.4 (2.8) |

BMI, body mass index

The primary objective of this study was to test for noninferiority of the new medical device AM-301 against an established comparator device (Nasaleze) containing HPMC. The mean TNSS (0-240 min) was 5.91 (SD = 1.45) during unprotected exposure (**Table 2**). It reduced to 4.82 (SD = 1.74) for subjects treated with AM-301 prior to the challenge and to 5.20 (SD = 1.70) for subjects treated with HPMC.

**Table 2.** Mean TNSS values after the allergen challenge while unprotected and while under AM-301 and HPMC treatment

|  | TNSS, mean (SD) [95% CI] |  |  |
| --- | --- | --- | --- |
|  | Unprotected | AM-301 | HPMC |
| Overall | 5.91 (1.45) [5.41, 6.40] | 4.82 (1.74) [4.23, 5.42]<sup>1</sup> | 5.20 (1.70) [4.62, 5.77]<sup>4</sup> |
| Sequence A | 5.52 (1.24) [4.90, 6.13] | 4.59 (1.67) [3.76, 5.42]<sup>2</sup> | 4.92 (1.47) [4.19, 5.65]<sup>2</sup> |
| Sequence B | 6.29 (1.58) [5.51, 7.08] | 5.07 (1.83) [4.13, 6.02]<sup>3</sup> | 5.47 (1.92) [4.52, 6.42]<sup>2</sup> |

<sup>1</sup> n = 35; <sup>2</sup> n = 18; <sup>3</sup> n = 17; <sup>4</sup> n = 36.

For the primary endpoint, least square means (LS_Means_) of the mean change in TNSS during challenge were estimated to be 4.74 and 5.14 under AM-301 and HPMC treatment, respectively, for a difference of -0.39 (95% CI, -0.89 to 0.10; p = 0.114) (**Table 3**). As the upper bound of the CI was less than the noninferiority margin of 1.2 points, noninferiority of AM-301 to HPMC was established. The coefficients of the fixed effect parameters were not significant for the sequence (p = 0.356) or the period (p = 0.795). Hence, there were no signs of a difference between sequences or periods.

**Table 3.** Differences in mean TNSS over 240 min during pollen challenge between AM-301 and HPMC, for the intention-to-treat population

| Mean TNSS |  | LS<sub>Mean</sub> | Standard error | 90% CI | 95% CI |
| --- | --- | --- | --- | --- | --- |
| Treatment LS<sub>Means</sub> |  |  |  |  |  |
|  | AM-301 | 4.75 | 0.291 | [4.26, 5.23] | [4.16, 5.33] |
|  | HPMC | 5.14 | 0.289 | [4.66, 5.62] | [4.56, 5.72] |
| Difference in LS<sub>Means</sub> |  |  |  |  |  |
|  | AM-301 - HPMC | -0.39 | 0.242 | [-0.80, 0.02] | [-0.89, 0.10] |

In the predefined exploratory analysis of secondary endpoints, the increase in mean TNSS from baseline was lower under AM-301 than HPMC treatment at all time points up to 240 min, except for 120 min (**Figure 3**). The difference was significant for AM-301 at 20 min (p = 0.013) and 40 min (p = 0.003) with an LS_Mean_ of -1.10 (95% CI, -1.95 to -0.25) and -1.15 (95% CI, -1.86 to -0.43), respectively. Treatment with either AM-301 or HPMC resulted in significantly lower increases in mean TNSS from the baseline than during the unprotected screening ACC visit, with 95% CIs excluding zero and p <0.0001: For AM-301, LS_Mean_ = -1.09 (95% CI, -1.65 to -0.54), and for HPMC, LS_Mean_ = -0.71 (95% CI, -1.20 to -0.22). The estimated difference was nominally larger under AM-301 than HPMC treatment. The protective effects of the two barrier-forming products persisted for the duration of the challenge, i.e. for 4 hours.

**Figure 3.**
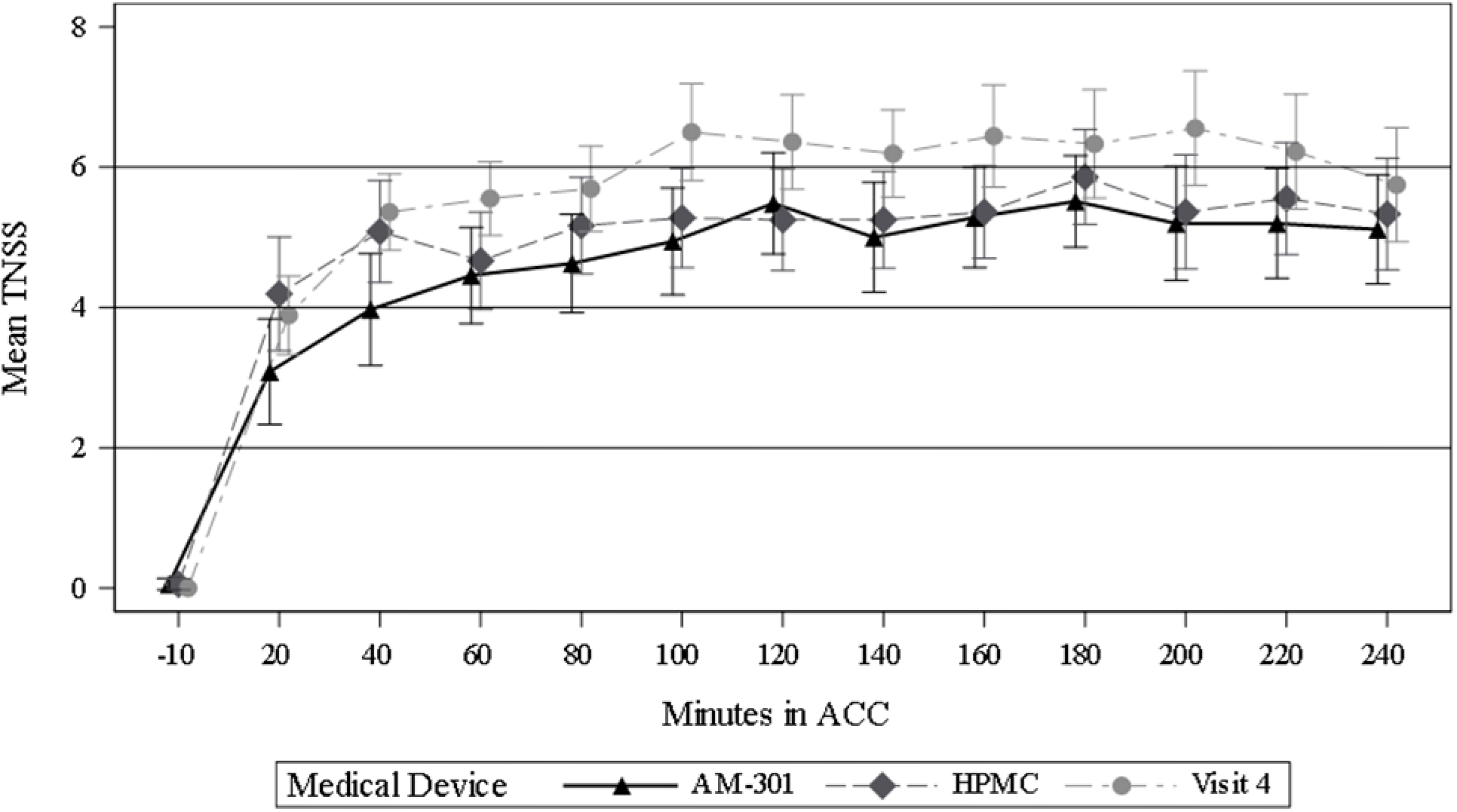
Mean values of the total nasal symptom score (TNSS) over time during the unprotected screening ACC visit and during medical device treatments (AM-301 and HPMC). Visit 4 corresponds to unprotected ACC visit. Error bars represent 95% confidence Intervals. ACC, allergen challenge chamber; Mean TNSS – averaged over subjects

When the TNSS results were categorized (severe, 9-12; moderate, 5-8; mild, 1-4; none, 0), severe and moderate symptoms were experienced less frequently under treatment with both devices than during unprotected exposure (**Figure 4**). At 20 and 40 min, AM-301-treated subjects had a higher percentage of mild symptoms (66% and 63%) than HPMC-treated subjects (44% and 42%).

**Figure 4.**
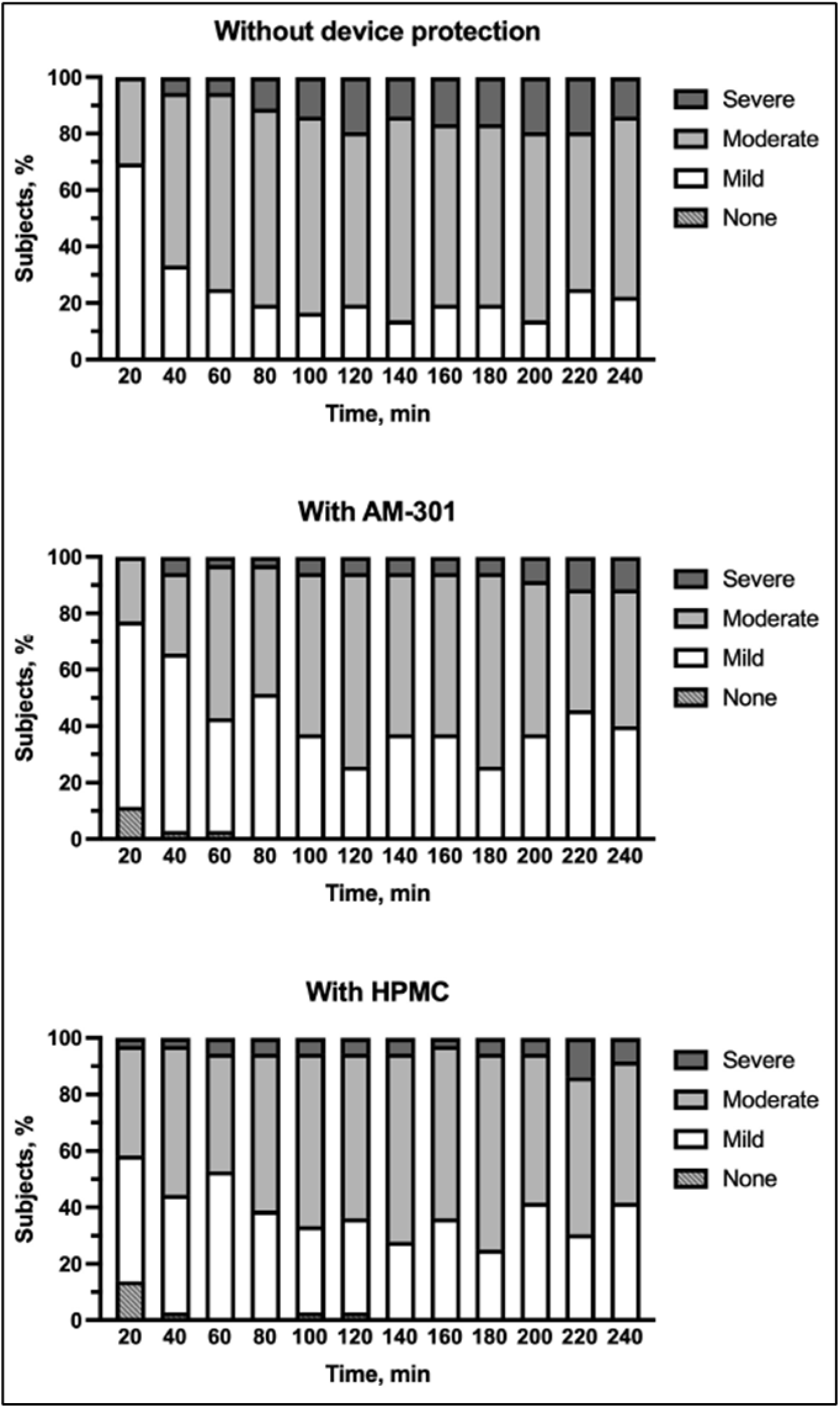
Categorized total nasal symptom scores over time during the unprotected screening ACC visit (“without device protection”, top) and during the treatment ACC visits with either AM-301 (middle) or HPMC (bottom) protection. Severe, 9-12; moderate, 5-8; mild, 1-4; none,0

Changes in mean TNSS item scores from unprotected exposure to device-protected exposure are shown in **Table 4**. Negative changes, indicating a reduction in score, were observed for every item after both treatments except for item sneezing after HPMC treatment. The absolute values of the changes were larger for AM-301 than HPMC. For AM-301, the 95% CIs excluded 0 for nasal congestion, rhinorrhea and nasal itching; for HPMC, the 95% CIs excluded 0 for nasal congestion and nasal itching. The treatment effect for rhinorrhea was significant (p = 0.04) with an LS_Mean_ of -0.15 (95% CI, -0.29 to -0.01), meaning that the decrease was significantly larger for AM-301 than for HPMC.

**Table 4.** Changes in means of individual TNSS items between device-protected and unprotected grass pollen exposures

| TNSS item | Change in mean of TNSS item (SD) [95% CI] |  |
| --- | --- | --- |
|  | AM-301 | HPMC |
| ITT | N=35 | N=36 |
| Nasal congestion | -0.33 (0.65) [-0.56, -0.11] | -0.27 (0.54) [-0.45, -0.08] |
| Rhinorrhea | -0.24 (0.47) [-0.40, -0.07] | -0.09 (0.47) [-0.25, 0.07] |
| Nasal itching | -0.46 (0.57) [-0.66, -0.27] | -0.34 (0.54) [-0.53, -0.16] |
| Sneezing | -0.06 (0.46) [-0.22, 0.10] | 0.00 (0.50) [-0.17, 0.16] |

The mean weight of nasal secretions during the unprotected visit was 14.5 g (SD = 13.9 g). This weight increased during the challenge to 17.8 g (SD = 17.6 g) for AM-301 and to 18.9 g (SD = 16.6 g) for HPMC. Although the weight of nasal secretions tended to be lower under AM-301 than HPMC treatment, the difference between treatments was not significant (p = 0.31). The period effect reached formal significance (p = 0.01), but no sequence effect was detected.

The efficacy of AM-301 on a 4-point scale was rated good or very good by 11 subjects (31%) and by the investigator for 16 cases (45%) (**Figure 5**). In contrast, HPMC was rated good or very good by 5 subjects (14%) and by the investigator for 9 cases (25%). Tolerability was rated as good or very good by over 80% of the subjects for both devices.

**Figure 5.**
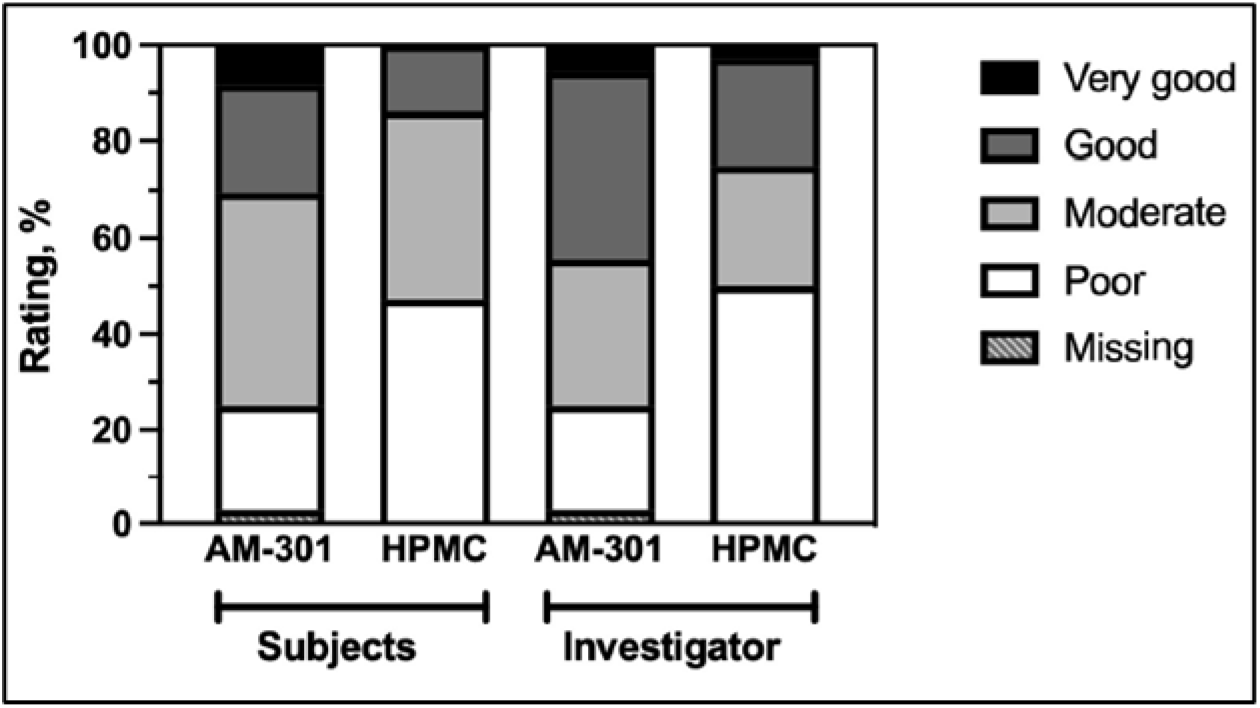
Overall subjective rating of efficacy by subjects and investigators. One rating for AM-301 is missing due to drop-out

A total of 48 adverse events were reported by 26 (72%) subjects. The intensity was mild for 36 (75%) and moderate for 12 (25%) adverse events. No serious adverse events occurred. After randomization, 34 adverse events were reported (25 after AM-301 and 9 after HPMC administration), and 18 of these (in 16 subjects) were considered medical device-related by the investigators. The differences between treatment groups reflected a short-lasting, burning sensation in the nose experienced by 15 subjects after administration of AM-301. None of the adverse events led to study withdrawal or were related to device deficiencies.

The study subjects had a mean blood pressure (systolic / diastolic) of 122.0 / 72.5 mmHg at the first visit of the screening period. Their mean heart rate was 66.6 beats/min. No clinically significant changes in these parameters were registered during the clinical investigation. PEF monitoring did not reveal any clinically relevant reduction in lung function after the allergen challenge for all randomized subjects. There was no significant difference in any study endpoint between the ITT and per-protocol population.

## Discussion

This clinical investigation compared the efficacy and safety of AM-301 to that of the comparator device Nasaleze Allergy Blocker (HPMC) in alleviating the symptoms of SAR triggered by grass pollen exposure in the validated Fraunhofer ACC. After a single-dose nasal application of either AM-301 or HPMC, efficacy was assessed via TNSS, nasal secretion weights, and subjective ratings.

AM-301 showed noninferiority towards HPMC in the primary endpoint and several exploratory endpoints. Both devices provided a significant reduction in overall symptoms compared to unprotected exposure. This finding confirms that HPMC is protective and indicates that AM-301 can protect at least to the same extent as HPMC. In the exploratory analysis of secondary endpoints, AM-301 gave significantly better protection at early time points (20 and 40 min) and for the TNSS item rhinorrhea. The faster onset of protection for AM-301 than HPMC may be due to differences in their physical states: AM-301 forms a gel immediately upon contact with the nasal mucosa, while HPMC is a powder that needs to swell within the nasal cavity to form the barrier.

The patient-reported efficacy assessments suggest that AM-301 could be better in reducing symptoms than the comparator device, which was corroborated by the investigators’ assessments. In particular, there was a large difference in the frequency of “poor” ratings, with more than twice as many HPCM-treated subjects selecting this category (48.6%) than AM-301-treated subjects (22.9%). Both subjects and the investigators rated the global efficacy of AM-301 more favorably than HPMC. Given that AM-301 is a new product, the subjects could not have had any preconceived expectations about it.

The data indicating a period effect for nasal secretion were unexpected. An increase due to repeated pollen exposure challenges can be excluded based on data from a validation study [15] and several other studies conducted with the Fraunhofer ACC [24,25]. The potential impact of the approaching allergy season was controlled by the criterion of TNSS ≤3 prior to the ACC visits, and subjects did not report symptoms corresponding to their grass pollen allergy when asked prior to the challenges. However, a seasonal impact cannot be fully excluded. The increase in nasal secretion with treatment compared to unprotected exposure occurred in both treatment groups to a similar degree. It is conceivable that topical application of the medical device increased the amount of fluid collected during the treatment ACC visits. Nonetheless, a treatment effect was not observed, and standard deviations for nasal secretion weight suggest a wide distribution of the data. Furthermore, increased nasal secretion weight does not correspond to the lower ratings for the TNSS item rhinorrhea after treatment with AM-301.

Overall tolerability favored HPMC over AM-301 due to the brief burning sensation experienced by several subjects. This effect may be attributed to the presence of preservatives (sodium benzoate, phenoxyethanol and methylparaben) in the formulation of AM-301 tested in this study. Given that people with allergic rhinitis are sensitive to nasal irritants, all preservatives have been eliminated from the formulation of the marketed version of AM-301.

Overall, the new medical device AM-301 consistently showed noninferiority towards HPMC across efficacy endpoints with a tendency for better performance. Furthermore, AM-301 provided more pronounced protection within in the first 40 min of allergen challenge than did HPMC treatment. Both barrier-forming products had a persisting protective effect over the 4-hour challenge, and both were safe to use. Altogether, the risk-benefit assessment of AM-301 is positive. Patients with SAR may benefit from its use for protection against airborne allergens.

## Supporting information

CONSORT Checklist

## Data Availability

Raw data are available from the corresponding author upon request. The medical devices used in the study are commercially available in some countries.

## Abbreviations

ACC: allergen challenge chamber
CI: confidence interval
HPMC: hydroxypropyl methylcellulose
ITEM: Institute for Toxicology and Experimental Medicine
LSmeans: least square means
PEF: peak expiratory flow
SAR: seasonal allergic rhinitis
SD: standard deviation
TNSS: total nasal symptom score

## Declarations

### Ethics approval and consent to participate

The study protocol was approved by the Ethics Committee of Hannover Medical School prior to study start. The trial was registered in the German Clinical Trial Register (DRKS00024356) and with EUDAMED (CIV-20-10-034870).

### Consent for publication

All subjects gave consent to the scientific publication of the data collected in this study.

### Authors’ contributions

JN and NG managed the study and drafted the manuscript. FF contributed to study design and conduct regarding the use of AM-301. IPH and PB gave major input to study design and had an oversight role in this study. JMH, PB, TM, IPH and FF revised the manuscript and gave scientific input to the study.

## Acknowledgments

Valerie Matarese provided scientific editing services for this manuscript. Michael Swora of X-act Cologne Clinical Research GmbH (Cologne, Germany) conducted the statistical analysis and provided feedback on the manuscript.

